# Sex workers’ experiences of HIV prevention, healthcare access and mental health in the West Rand District, South Africa: a qualitative study

**DOI:** 10.64898/2026.10.13.26365391

**Authors:** Zwonaka Maphauphau, Moreoagae Bertha Randa, Muhammad Hoque

## Abstract

Sex workers experience disproportionate HIV vulnerability, while access to prevention, healthcare and mental health support is shaped by economic, social and institutional conditions. This study explored sex workers’ experiences of HIV prevention, healthcare access and mental health in the West Rand District, South Africa. A qualitative descriptive-interpretive design was used. Twenty unique participants were recruited through purposive and snowball sampling: five completed individual semi-structured interviews and 15 different participants took part in two focus group discussions comprising seven and eight participants. Data were audio-recorded with permission, transcribed verbatim and analysed using reflexive thematic analysis informed by Braun and Clarke’s six-phase approach, with NVivo 15 used for data organisation. Six themes were developed: sex work as economic survival rather than simple choice; HIV prevention under unequal client power; healthcare access shaped by stigma, fear and confidentiality concerns; participant-reported mental health distress as part of everyday sex work; peer educators as trusted bridges to care; and the need for inclusive and differentiated support. Participants described substantial knowledge of condoms, HIV testing and pre-exposure prophylaxis, but their accounts indicated that prevention preferences were negotiated within income needs, client demands, safety concerns and disclosure risks. Participants also reported anxiety, stress, trauma-related experiences, depressive symptoms and substance use in relation to work and healthcare experiences; these reports were not clinical diagnoses. Peer educators were described as trusted sources of information, referral, emotional support and clinic navigation. Overall, the findings suggest that meaningful access extends beyond geographical availability and is shaped by confidentiality, dignity, economic opportunity costs and trusted relationships. The findings support consideration of integrated, differentiated and peer-linked approaches in district-level HIV, healthcare and mental health planning.

## INTRODUCTION

Sex workers remain a priority population in the global and South African HIV response because HIV vulnerability is produced through interacting social, economic, legal and institutional conditions rather than individual behaviour alone. The World Health Organization (2022) identifies sex workers as a key population requiring comprehensive, voluntary and acceptable HIV, sexually transmitted infection and viral hepatitis prevention, diagnosis, treatment and care. UNAIDS (2024) similarly emphasises that stigma, violence, discrimination and punitive legal environments continue to restrict sustained access to prevention and treatment. In South Africa, the National Strategic Plan for HIV, TB and STIs 2023-2028 promotes differentiated, rights-based and community-centred services for key populations (South African National AIDS Council, 2023). Earlier global evidence also shows that HIV outcomes among sex workers are inseparable from structural conditions affecting safety, rights and healthcare access (Shannon et al., 2018). Meaningful HIV prevention access must therefore be understood as the capacity to obtain and use services safely, consistently and with dignity, rather than as the simple presence of condoms, testing or medication.

HIV prevention choices are made within working environments characterised by variable income, mobility, client bargaining power, violence and stigma. South African epidemiological evidence confirms a substantial burden of HIV among female sex workers and underlines the continuing need for effective prevention and treatment strategies (Kassanjee et al., 2022). Oral pre-exposure prophylaxis (PrEP) has expanded prevention options, yet its uptake and continued use are shaped by individual motivations, disclosure concerns and the social meanings attached to antiretroviral medicines (Makhakhe, Meyer-Weitz and Sliep, 2022). Programme evidence further indicates that implementation strategies such as provider mentoring and reminder support can affect PrEP persistence, although persistence remains challenging in real-world service delivery (Rao et al., 2023). These findings indicate that prevention knowledge and prevention agency are not equivalent. A sex worker may understand available prevention methods while still negotiating their use within relationships in which clients control payment, location or the terms of an encounter.

The relationship between psychological well-being and HIV prevention is also interconnected. Female sex workers living with HIV in eThekwini have been reported to experience depression and other syndemic factors associated with poorer HIV outcomes (Bhardwaj et al., 2023). Research in Soweto and Klerksdorp has likewise documented substantial alcohol and drug use among female sex workers and links between substance use, trauma and other vulnerabilities (Yeo et al., 2022). At service-system level, evidence from HIV programmes suggests that integrating mental health care with HIV services can support engagement and psychological outcomes, although approaches and effectiveness vary by context (Conteh, Latona and Mahomed, 2023). A recent systematic review similarly identifies potential value in integrated HIV and mental health service delivery in low- and middle-income countries (Azuma et al., 2025). South Africa’s National Mental Health Policy Framework and Strategic Plan 2023-2030 further supports integration of mental healthcare into community and district health services (National Department of Health, 2023). For the present study, mental health is therefore treated as part of the broader service environment rather than as an isolated clinical outcome.

Healthcare access also involves privacy, dignity, trust, safety and continuity of care. Chen et al. (2022) found that HIV-related and sex work-related stigma was associated with poorer quality of life among female sex workers living with HIV in South Africa. Comins et al. (2024) showed that treatment coverage and viral suppression among female sex workers are shaped by both individual and structural vulnerabilities, demonstrating the importance of how services are organised. International guidance similarly stresses acceptable and non-discriminatory care for key populations (World Health Organization, 2022), while UNAIDS (2024) identifies stigma and fear of disclosure as continuing barriers to service engagement. Consequently, geographical proximity to a clinic cannot be assumed to represent meaningful access. Where attendance may reveal HIV status, expose sex work, or lead to anticipated judgement, individuals may conceal relevant information, delay care or disengage from follow-up even when services are physically available.

Rhodes’ Risk Environment Framework provides the conceptual basis for examining these interconnected experiences. The framework moves analysis away from individualised explanations of risk towards the physical, social, economic and policy or institutional environments in which health practices occur (Rhodes, 2002). In this study, the framework was used as an interpretive lens rather than as a deductive coding scheme. Initial coding remained primarily inductive, after which the developing themes were considered in relation to the framework’s environmental dimensions. This approach allowed the analysis to retain participants’ own meanings while examining how income insecurity, client power, workplace location, stigma, violence, confidentiality and service organisation shaped the scope for prevention and healthcare engagement. Peer educators were considered as relational actors operating across community and service environments.

### Problem Statement and Aim

Three related gaps motivate the study. First, South African research has established substantial HIV risk and has examined prevention options such as PrEP among sex workers, but less is known about how economic dependence and unequal client power shape the practical ability to implement prevention preferences in everyday encounters (Kassanjee et al., 2022). Evidence on PrEP motivation and persistence shows that prevention behaviour is responsive to personal and programme factors, yet it does not fully explain how immediate income needs and client bargaining influence condom use, disclosure and refusal of unsafe requests (Makhakhe, Meyer-Weitz and Sliep, 2022; Rao et al., 2023). This gap is important because a focus on knowledge or service availability alone may overstate the degree of control that individuals can exercise within economically unequal interactions.

Second, existing South African evidence clearly identifies HIV-related and sex work-related stigma, but fewer studies explain how anticipated confidentiality breaches, clinic visibility and healthcare-worker interactions shape engagement with services at district level. Chen et al. (2022) demonstrate the importance of stigma for quality of life, while Comins et al. (2024) show that structural vulnerabilities remain relevant to treatment outcomes. Third, mental health and HIV evidence often remains programme-specific, even though sex workers may experience emotional distress, substance use, client negotiation, violence and healthcare engagement as interconnected features of everyday life (Bhardwaj et al., 2023). Reviews of integrated care support closer attention to the relationship between mental health and HIV service delivery (Conteh, Latona and Mahomed, 2023; Azuma et al., 2025). The study therefore aimed to explore sex workers’ experiences of HIV prevention, healthcare access and mental health in the West Rand District of South Africa, with particular attention to how economic, social, physical and institutional environments shaped those experiences. The term “experiences” is used here to refer to participants’ first-person accounts and interpretations; the study was not designed as a phenomenological inquiry.

## METHODS

### Research Design

The study used a qualitative descriptive-interpretive design with reflexive thematic analysis to explore participants’ experiences, perceptions and interpretations of HIV prevention, healthcare access and mental health. A qualitative approach was appropriate because the research question concerned how sex workers made sense of economic pressures, client relationships, stigma, emotional distress, peer support and healthcare encounters within their social contexts. The manuscript uses the term “experiences” to describe participants’ first-person accounts and meanings rather than to claim a phenomenological methodology. Reflexive thematic analysis was selected because themes are understood as analytic constructions developed through the researcher’s engagement with the dataset rather than as objective entities waiting to be discovered. The study was prepared with reference to the Consolidated Criteria for Reporting Qualitative Research and the Standards for Reporting Qualitative Research to strengthen transparency regarding the research team, setting, recruitment, data generation, analysis and interpretation.

### Setting

The study was conducted in the West Rand District of Gauteng Province, South Africa. The district contains urban, peri-urban and informal-settlement communities shaped by mining activity, unemployment, population mobility and uneven access to public healthcare. Participants described sex work occurring across streets, entertainment venues, lodges, private residences, client-arranged locations and online or privately negotiated settings. These environments differ in visibility, exposure to potential violence and contact with outreach services. The West Rand was therefore an appropriate setting for examining how local economic conditions, transport requirements, clinic organisation, social stigma and safety concerns intersect with HIV prevention and mental health support. Providing this contextual description also assists readers in judging whether the findings may be transferable to comparable district-level settings rather than assuming that experiences in the West Rand represent all sex workers in South Africa.

### Study Population and Eligibility Criteria

The study population comprised adults engaged in sex work who lived or worked in the West Rand District. Participants were eligible if they were 18 years or older, self-identified as sex workers and had experience or knowledge of HIV prevention services, mental health support, general healthcare or peer-led interventions within the district. Participation was not restricted by gender identity, sexual orientation, HIV status, antiretroviral treatment use or enrolment in a specific healthcare programme. Individuals younger than 18 years, those who were not currently engaged in sex work and those who neither lived nor worked in the district were excluded. These criteria ensured that participants could provide current, first-hand accounts of the service environments examined in the study while permitting diversity in work arrangements and personal identities.

### Participant Sampling and Recruitment

Purposive sampling was initially used to identify sex workers with experience of HIV prevention, healthcare and peer-linked services. Snowball sampling was subsequently used to extend recruitment to individuals who were less visible, including people working through private or online arrangements or those not routinely connected to formal healthcare. Recruitment was supported through peer educators, community-based organisations, outreach activities and existing service contact points. These intermediaries shared information about the study, while interested individuals contacted the research team directly rather than being selected by peer educators. This approach was intended to reduce pressure to participate and to limit unnecessary disclosure of participation within peer networks. However, recruitment through peer and service networks may have favoured participants who were already connected to outreach or healthcare and may have underrepresented people who were more isolated from such networks. This potential selection effect is considered in the limitations.

The final sample comprised 20 unique participants. Five completed individual semi-structured interviews, while 15 different participants took part in two focus group discussions, with seven participants in the first group and eight in the second. Before data collection, participants received information about the study purpose, procedures, potential risks and anticipated benefits and provided written informed consent. Participation was voluntary, and participants could decline to answer any question, request a break or withdraw without losing access to healthcare or peer-led services. Participants were recruited and participated between 19 and 25 March 2026. No individual interview participant attended a focus group, and each participant contributed to one data-collection activity. Recruitment and preliminary analytic engagement occurred iteratively, allowing the research team to assess the breadth and depth of the developing dataset. Because recruitment was partly facilitated through peer and outreach networks, the possibility that people already connected to services were more likely to be reached was recognised and is considered in the limitations.

### Sample Adequacy and Thematic Depth

The sample size was not justified by treating saturation as an objective endpoint. Instead, adequacy was considered in relation to the specificity of the research question, the focused participant group, the complementary use of individual interviews and focus groups, the richness of the accounts, variation in work settings and identities, and iterative engagement with the data during collection and analysis. Hennink and Kaiser (2022) provide empirical guidance on sample adequacy in qualitative research, but reflexive thematic analysis requires caution in presenting saturation as if themes exist independently of researcher interpretation. Recruitment concluded after the 20th participant because the dataset provided sufficient thematic depth to address the research aim and supported coherent interpretive development across the six themes. Later accounts continued to add contextual detail and variation; therefore, the decision to stop recruitment was based on overall analytic adequacy rather than a claim that no further meanings could be generated.

### Data Collection

Data were generated through five individual semi-structured interviews and two focus group discussions. Individual interviews provided a private format for discussing potentially sensitive experiences, including HIV status disclosure, client behaviour, clinic encounters and emotional distress. Focus groups were used to explore shared working conditions, healthcare barriers, peer support and service experiences that participants were comfortable discussing collectively. Given the sensitivity of the topic, facilitators avoided requesting disclosure of identifiable HIV status or other information that participants did not wish to discuss publicly, and participants were reminded that they should not repeat other participants’ contributions outside the group. The interview and focus-group guides addressed sex work in the West Rand, condom negotiation, PrEP, antiretroviral treatment, HIV testing, sexually transmitted infection care, healthcare stigma, confidentiality, mental health experiences, coping strategies, peer-led interventions and suggestions for improving services. Each participant took part once, and no individual interview participant was subsequently included in a focus group.

Data collection took place in private, secure and participant-accessible locations, including healthcare facilities, drop-in centres, mobile outreach sites and peer-identified safe spaces. Individual interviews lasted approximately 20–30 minutes, and each focus group was approximately 90 minutes. The individual interviews were relatively focused encounters, and their shorter duration is acknowledged as a limitation when interpreting the depth of individual narratives. Sessions were conducted in English or another language selected by the participant. Where a language other than English was used, the transcript was translated into English and cross-checked against the audio recording to preserve meaning. All sessions were audio-recorded with permission, and field notes documented contextual observations, group interaction, non-verbal communication and preliminary analytic reflections. The first author facilitated all individual interviews and focus groups and was identified to participants as a postgraduate public health researcher rather than as a healthcare provider, law-enforcement official or service gatekeeper.

### Data Analysis

The data were analysed using reflexive thematic analysis informed by Braun and Clarke’s six-phase approach. NVivo 15 was used to organise transcripts, codes, analytic notes and quotations, but analytic decisions remained the responsibility of the research team. The first author began by listening to the recordings, repeatedly reading transcripts and reviewing field notes to become familiar with the dataset. Initial observations focused on income insecurity, condom negotiation, healthcare encounters, emotional distress, coping, peer support and differences across work settings and identities. The transcripts were then coded systematically. Coding was primarily inductive, while remaining sensitised by the research aim and Rhodes’ Risk Environment Framework. The framework was applied during theme development and interpretation rather than used to force data into predefined categories. This balance allowed unexpected meanings to remain visible while also examining how economic, social, physical and institutional conditions appeared in participants’ accounts.

Related codes were developed into candidate themes representing patterned meanings across interviews and focus groups. Candidate themes were reviewed against coded extracts and the wider dataset to assess their coherence, distinctiveness and relevance to the research aim. Divergent accounts were retained rather than treated as errors, because reflexive thematic analysis does not assume a single uniform experience among sex workers. Themes were subsequently defined and named to clarify their central organising concepts, and representative quotations were selected to illustrate the interpretation. The first author conducted primary coding and theme development. Co-authors reviewed the developing analytic framework, challenged interpretations, considered alternative explanations and discussed the coherence and boundaries of themes. These discussions supported reflexive interpretation and analytic depth; they were not used as an inter-rater reliability exercise or as a search for one objectively correct coding solution.

### Trustworthiness

Credibility was supported by using both individual interviews and focus group discussions, enabling comparison between private accounts and collectively discussed experiences. Audio recordings were transcribed verbatim and checked against the recordings, while field notes preserved contextual information that could not be fully represented in transcripts. Formal participant member checking was not conducted. Instead, the analysis relied on repeated engagement with the dataset, transcript checking, reflexive analytic discussion, attention to divergent accounts and close grounding of interpretations in participant quotations. Dependability was supported through an audit trail containing recruitment decisions, interview and focus-group guides, field notes, coding decisions, analytic notes and theme development. Confirmability was strengthened through reflexive documentation and co-author challenge of developing interpretations, while transferability was supported through detailed reporting of the West Rand setting, partpathways, participant diversity, sex-work environments and healthcare context.

### Reflexivity

Researcher positionality was considered throughout data generation and analysis because discussions of sex work, HIV, stigma and mental health can be shaped by perceived professional authority and assumptions about risk or healthcare behaviour. The first author entered the field as a postgraduate public health researcher rather than as a healthcare provider, law-enforcement official or service gatekeeper. Participants were informed of the researcher’s role, the purpose of the study and the voluntary nature of participation, including that declining participation would not affect access to healthcare or peer-led services. Reflexive notes were written after interviews and focus groups to document assumptions, emotional responses, power dynamics and possible influences on questioning and interpretation. Particular attention was given to avoiding moral assumptions about sex work, HIV status, substance use, condom negotiation and clinic attendance. Co-authors challenged emerging interpretations and encouraged consideration of alternative explanations, but were not used as independent coders or to calculate inter-rater reliability.

Reflexivity also informed interpretation of diversity within the sample. The analysis did not assume that women, men, transgender women or participants identifying as bisexual experienced risk, stigma or healthcare access in the same way. Equally, subgroup differences were not asserted when the dataset did not provide sufficiently detailed evidence to support them. Interpretations were repeatedly checked against participant accounts, and co-authors provided methodological and interpretive challenge to encourage consideration of alternative explanations. The analysis therefore acknowledges that findings were produced through interaction between participants, the first author’s questioning and the research team’s interpretive work. This approach is consistent with reflexive thematic analysis, in which researcher subjectivity is treated as an analytic resource to be examined rather than an influence that can be eliminated entirely (Braun and Clarke, 2021).

### Ethical Considerations

Ethical approval was obtained from the Sefako Makgatho Health Sciences University Research Ethics Committee (SMUREC) under approval number SMUREC/H/544/2025:PG on 21 November 2025. Permission to conduct the study was also obtained from the West Rand Health District under reference GP_202510_091 and from relevant organisational gatekeepers. Participants received reimbursement in accordance with the approved study procedures; it was explained as reimbursement rather than an inducement, and participants retained it if they withdrew after data collection had commenced.

Participant names were not recorded in transcripts or reports. Unique study codes were used for individual interviews and focus groups, and identifying details were removed during transcription and quotation selection. Digital recordings, transcripts and NVivo files were stored on password-protected devices accessible only to the authorized research team. Focus-group participants were asked to respect the privacy of information shared by others and were explicitly informed that complete confidentiality between group members could not be guaranteed. Data-collection locations were selected to minimise the possibility of participants being identified as sex workers or service users. Because discussions could evoke distress associated with violence, HIV, stigma or mental health difficulties, the interviewer monitored emotional responses and offered pauses or termination of the session where necessary. These procedures sought to reduce disclosure and distress risks while recognizing the confidentiality challenges created by focus-group research with a small, potentially identifiable population.

## RESULTS

### Participant Characteristics

The qualitative sample comprised 20 unique sex workers. Five participants completed individual interviews, and 15 different participants took part in the two focus groups. Participants were aged 21-42 years and lived or worked in the West Rand District. The sample included women, men and transgender women, and some participants identified as bisexual. Gender identity and sexual orientation were treated as separate dimensions rather than being forced into mutually exclusive categories. Sex-work arrangements included street-based, venue-based, online or private and mixed forms of work. To reduce the risk of deductive disclosure in this small and potentially identifiable population, Table 1 reports only broad characteristics relevant to interpretation and omits detailed combinations of gender, HIV status, education and other attributes. This approach preserves the diversity of the sample without presenting combinations that could enable recognition of individual participants.

**Table 1:** Characteristics of the qualitative participants.

| Characteristic | Study sample |
| --- | --- |
| Total participants | 20 |
| Individual interviews | 5 |
| Focus group participants | 15 |
| Age | 21–42 years |
| Gender identities | Women, men and transgender women |
| Sex-work settings | Street-based, venue-based, online/private and mixed |
| HIV status | Positive and negative |
| Years engaged in sex work | 3–12 years |

### Overview of Themes

Six themes were developed from individual interviews and focus-group discussions. Participants described sex work as closely connected to economic survival, while HIV prevention was negotiated within unequal relationships with clients. They described healthcare experiences shaped by stigma, fear and confidentiality concerns and reported anxiety, stress, trauma-related experiences, depressive symptoms and substance use in relation to everyday work and healthcare encounters. Peer educators were described as trusted intermediaries who provided information, referrals, emotional support and clinic navigation. Participants also described varied work arrangements and identities, highlighting the importance of avoiding a single assumed profile of a sex worker. Across the themes, the accounts suggested that prevention knowledge did not automatically translate into prevention agency and that service availability did not guarantee meaningful access; economic pressures, client relationships, perceived safety and institutional trust remained important parts of the context in which decisions were made.

### Theme 1: Sex Work as Economic Survival Rather Than Simple Choice

Sex work was predominantly described as a livelihood strategy shaped by unemployment, poverty, family responsibilities and limited access to alternative income. Participants did not present their work solely as an unrestricted personal choice. Their accounts indicated that sex work provided immediate income where formal employment was unavailable, unstable or insufficient to meet household needs. Participants also linked changes in demand and prices to wider economic conditions in the West Rand, including reduced mining activity, unemployment and competition for clients. Income was described as unpredictable and dependent on client availability, location and daily circumstances. Within these accounts, decisions concerning health, safety and work were made alongside an immediate need to secure money for food, transport and family responsibilities. Economic conditions were therefore described as part of the setting within which prevention preferences and healthcare decisions were negotiated.

> *“The business is not going well since many mines have closed down, so I do not get the customers like in the past … my prices are declining because I am no longer in demand … I am desperate for money, any amount I agree to it so that I can survive with my kids.”*
>
> ***(SWFG2, Participant 3)***

Participants described financial pressure as limiting how firmly they felt able to negotiate with clients. Some clients were said to use knowledge of participants’ need for money to request lower prices or to change previously agreed terms. Participants linked the possibility of losing income with decisions about whether to accept a client, travel to a particular location or insist on preferred safety practices. At the same time, participants asserted sex work as work, labour and business and did not describe themselves only as passive victims. They spoke about prices, clients, demand and working hours in ways that reflected the organisation of income-generating activity. These accounts show the coexistence of agency and constraint: participants described active decision-making, but within circumstances in which income insecurity could narrow the range of options that felt practically available.

> *“You are not just a free agent. You are still working. You are still somebody who is working. So it is very important that you take care of yourself, your mental health, and your physical health.”*
>
> ***(SWI-P2)***

Participants also connected clinic attendance with potential opportunity costs. Time spent travelling to a facility, waiting for medication or attending repeated appointments could coincide with periods in which income might otherwise be earned. Similar concerns arose when refusing clients or leaving an unsafe encounter. The accounts therefore positioned healthcare use and prevention behaviour within daily livelihood decisions rather than as separate health choices. Participants did not describe these opportunity costs as affecting every encounter in the same way, but they repeatedly appeared in explanations of why preferred health practices could be difficult to maintain. This finding is reported here as an experience described by participants rather than as a measured causal relationship between poverty and HIV prevention behaviour.

### Theme 2: HIV Prevention Under Conditions of Unequal Client Power

Participants generally demonstrated knowledge of condoms, HIV testing, PrEP, antiretroviral treatment and sexually transmitted infection prevention. Knowledge, however, did not consistently correspond with the ability to apply preferred prevention practices in every encounter. Participants described negotiations in which clients-controlled payment, location or other terms of the interaction. Client resistance to condoms or offers of additional payment for sex without protection could create circumstances in which economic need conflicted with participants’ preferred prevention choices. These accounts suggest a distinction between knowing how HIV can be prevented and having sufficient relational and economic power to act on that knowledge. The qualitative data does not establish that client power caused outcomes, but participants repeatedly located prevention decisions within negotiations over money, safety and the continuation of the encounter.

> *“How they pay determines what type of services they want … services with protection have a different price and services without protection also have a different price, so the moment they pay, that is when you will know if they want it with protection or not.”*
>
> ***(SWFG2, Participant 6)***

Participants also described HIV-status disclosure as a complex decision shaped by anticipated consequences. Before disclosing, participants considered possible rejection, loss of income, stigma, privacy and client reactions. Some described assumptions by clients that sex workers were living with HIV, while others noted that a client’s claimed HIV status could enter negotiations about condom use. Disclosure was therefore presented as a situated decision rather than a simple matter of knowledge or personal responsibility. At the same time, participants described forms of prevention agency. Some reported refusing sex without condoms, testing regularly or using PrEP as an additional prevention method. Peer educators were frequently mentioned as sources of understandable information about prevention options, particularly where clinic-based communication was experienced as judgemental or difficult to navigate.

> *“Yes, I am on PrEP and I have been taking it for some time now. It was because the peer educators told me all about it and I became more interested in knowing how it is used and how I could use it.”*
>
> ***(SWFG2, Participant 8)***

Taken together, participants’ accounts revealed a recurring tension between prevention knowledge and prevention agency. The data did not suggest that limited use of a prevention method was necessarily the result of ignorance. Rather, participants described circumstances in which client demands, financial pressure, safety concerns and disclosure risks affected the options they considered feasible. The theme therefore captures the practical negotiation of prevention within unequal relationships without claiming a deterministic effect of economic or client power on behaviour. This distinction is important because the findings are based on participants’ interpretations of their experiences rather than on experimental or longitudinal evidence of causality.

### Theme 3: Healthcare Access Shaped by Stigma, Fear and Confidentiality Concerns

Healthcare facilities were sometimes experienced as emotionally unsafe because participants anticipated judgement, unwanted visibility and weak confidentiality. Smaller clinics were described as particularly sensitive where community members could recognise the purpose of particular rooms, queues or service points. Participants expressed concern that being seen entering an HIV testing room or collecting medication could expose their health status. Others described healthcare-worker interactions in which disclosure of sex work appeared to prompt assumptions about sexually transmitted infections or personal responsibility for illness. Participants associated harsh communication, judgement after missed appointments and inadequate privacy with reduced trust in formal services. These accounts indicate that a clinic could be geographically accessible while still being experienced as socially difficult to use when attendance or disclosure was perceived to carry a risk of stigma.

> *“The clinics are very small. So, a room where you are going in for HIV testing, everybody knows … if I come out there and the results are positive, now I am going to be stigmatised.”*
>
> ***(SWI-P3)***
>
> *“I prefer peer educators than nurses because to them when you tell them that you are a sex worker, they start to generalise the issue, saying you have STI because you are a sex worker, what did you expect?”*
>
> ***(SWFG2, Participant 3)***

Participants also described peer accompaniment as useful when formal healthcare settings felt intimidating or difficult to navigate. Peer educators were said to assist with explaining symptoms, understanding referrals, communicating with healthcare workers and completing clinic visits. One participant attributed receipt of care to the involvement of a peer educator who accompanied them and explained the concern to a nurse. The accounts therefore placed healthcare access within relationships of trust and communication rather than treating access as a matter of distance alone. Although participants represented different gender identities and sexual orientations, subgroup numbers were small and the available quotations did not support confident claims that specific experiences belonged uniquely to men, bisexual participants or transgender participants. The analysis therefore reports the broader pattern of anticipated stigma and confidentiality concerns without overextending subgroup comparisons.

> *“I explained to my peer educator, who then went to the clinic with me and explained to the nurse … that’s when I got help.”*
>
> ***(SWFG2, Participant 5)***

### Theme 4: Participant-Reported Mental Health Distress in Everyday Sex Work and Healthcare Experiences

Participants reported anxiety, stress, fear, trauma-related experiences, emotional exhaustion and depressive symptoms in relation to unpredictable clients, unsafe locations, non-payment, stigma and the possibility of violence. These terms represent participants’ accounts and are not presented as clinical diagnoses because the study did not employ diagnostic instruments or formal mental health assessments. Anxiety was described in anticipation of meeting unfamiliar clients, particularly when a client selected an isolated or distant location. Participants described having to assess potential danger while also considering the financial consequences of refusing an encounter. Participants also described distress in relation to healthcare experiences, including mistrust, anticipated judgement and difficult navigation of services. The findings therefore indicate that mental health was discussed as part of the same economic, relational and institutional environment in which HIV prevention and healthcare decisions occurred.

> *“I usually have anxiety especially when I have to meet a client for the first time, because it is scarier especially when they book a place far away from where you stay or where you are more comfortable.”*
>
> ***(SWFG2, Participant 4)***

Non-payment and changes to agreed terms were also described as sources of distress. Participants reported situations in which clients refused payment after an encounter or used payment to exert control. Such experiences were described in terms of both financial loss and emotional strain because participants had invested time, transport costs and personal safety without receiving the expected income. The expression ‘mental breakdown’ is retained only in the following participant quotation and is not used as a diagnostic label by the researchers. Participants’ accounts suggested that perceived inability to challenge a client safely could be accompanied by feelings of powerlessness, frustration or exhaustion. These findings describe how participants understood the emotional consequences of particular encounters; they do not establish a clinical disorder or a direct causal pathway from sex work to mental illness.

> *“I once had a mental breakdown … I went to a client’s place, and he did not want to pay … I just said okay and used my money to go back home.”*
>
> ***(SWFG2, Participant 3)***

Participants described varied ways of coping with distress. Some sought support from trusted friends, family members, experienced sex workers or peer educators, while others reported using alcohol or cannabis to manage anxiety, emotional pain or mistrust. Substance use was therefore discussed by participants as one coping strategy among several rather than being treated as a diagnosis or an inevitable feature of sex work. Some accounts also suggested that emotional exhaustion, fear and mistrust could make clinic attendance or prevention negotiation feel more difficult, although the qualitative design cannot determine the direction or magnitude of such relationships. The theme consequently positions participant-reported distress and coping within the same risk environment as client negotiation, violence, stigma and healthcare engagement.

> *“For me my coping mechanism is drinking alcohol and smoking weed … I have reached a point where I do not trust people anymore.”*
>
> ***(SWFG2, Participant 2)***

### Theme 5: Peer Educators as Trusted Bridges to Care

Peer educators were repeatedly described as trusted intermediaries between sex workers and formal healthcare services. Participants valued peers because they were perceived to understand the realities of sex work, communicate without moral judgement and remain accessible in spaces where participants worked or felt safe. Peer educators provided information about condoms, PrEP, HIV testing and treatment and were also described as sources of referrals, clinic accompaniment and emotional reassurance. Participants contrasted this relational accessibility with formal services that could feel impersonal or judgemental. The importance attributed to peers therefore extended beyond the transmission of health information. Participants described trust, familiarity and the way information was communicated as central to whether support felt usable.

> *“Peer-led interventions are bridging a gap between communities and the clinic … with the peer-led interventions, it is quite easier because now they are coming to us.”*
>
> ***(SWI-P3)***

Participants also described peers as assisting when clinic attendance or treatment engagement felt difficult. They reported encouragement to attend appointments, explanations of treatment requirements and help with arranging or navigating referrals. For some participants, accompaniment reduced uncertainty about formal services and made the process of seeking care feel less isolating. Emotional support formed part of this role. Participants described being able to discuss fears, stigma, client experiences and health concerns with peers when such conversations felt difficult elsewhere. These accounts position peer educators as boundary-spanning actors who connect community experience with institutional services. However, participants did not suggest that peers could replace all professional healthcare or mental health functions; rather, their accounts described peers as a trusted pathway into services and a source of support around formal care.

> *“The lady that was assisting me amongst the peer educator team kept on encouraging me to go to the clinic and to also take my medication … she made the first appointment for me and then when I got there, it was much easier.”*
>
> ***(SWI-P3)***
>
> *“With the peer educators, it becomes easier because now you feel like you have safe hands that you can always go to … to tell them about your problems, tell them about your fears.”*
>
> ***(SWI-P2)***

### Theme 6: Inclusive and Differentiated Support Across Diverse Work Settings

Participants’ accounts indicated that the sex-worker population in the West Rand was heterogeneous. The sample included women, men and transgender women, participants identifying as bisexual, and people working in street-based, venue-based, online or private and mixed settings. Work arrangements produced different levels of visibility and contact with outreach or clinic services. Street-based work was discussed in relation to public visibility and direct outreach contact, whereas online or private work could reduce routine contact with street-focused services. These observations do not establish systematic subgroup differences because the numbers within particular identity categories were small. Rather, they indicate that service reach and confidentiality needs may vary according to how and where people work.

Participants’ broader accounts supported the importance of service approaches that can respond to varied work arrangements, confidentiality needs and preferred modes of contact. Requests for privacy, non-judgemental communication, peer support and easier navigation appeared across the dataset, while the practical form of support could differ according to how and where participants worked. The theme is therefore interpreted as an empirical account of participants’ concerns about accessibility and fit, rather than as a claim that particular identity groups require fixed or separate services. It also highlights a potential gap in outreach where people are less visible to established street-based networks or anticipate disclosure and judgement in conventional services. Because this theme is closely linked to service implications, it should be read together with Themes 3 and 5 rather than as a standalone prescription for programme design.

## DISCUSSION

### Interpretation of the Main Findings

The study’s principal contribution is an account of how HIV prevention, healthcare access and participant-reported mental health distress were experienced within an interconnected risk environment. Rhodes’ Risk Environment Framework helps organise these experiences across economic, social and relational, physical, and policy or institutional dimensions (Rhodes, 2002).

Participants described unemployment, poverty and unstable income as part of the economic environment; client bargaining power, stigma and family responsibilities within the social and relational environment; isolated meeting locations and clinic visibility within the physical environment; and confidentiality, healthcare-worker judgement and service organisation within the institutional environment. Peer educators and trusted relationships operated as enabling features across these dimensions. Table 2 maps the empirical findings onto the framework, while Figure 1 presents an interpretive pathway linking these domains. The arrows represent relationships suggested by participants’ accounts and do not indicate causal effects established by this qualitative study.

**Figure 1.**
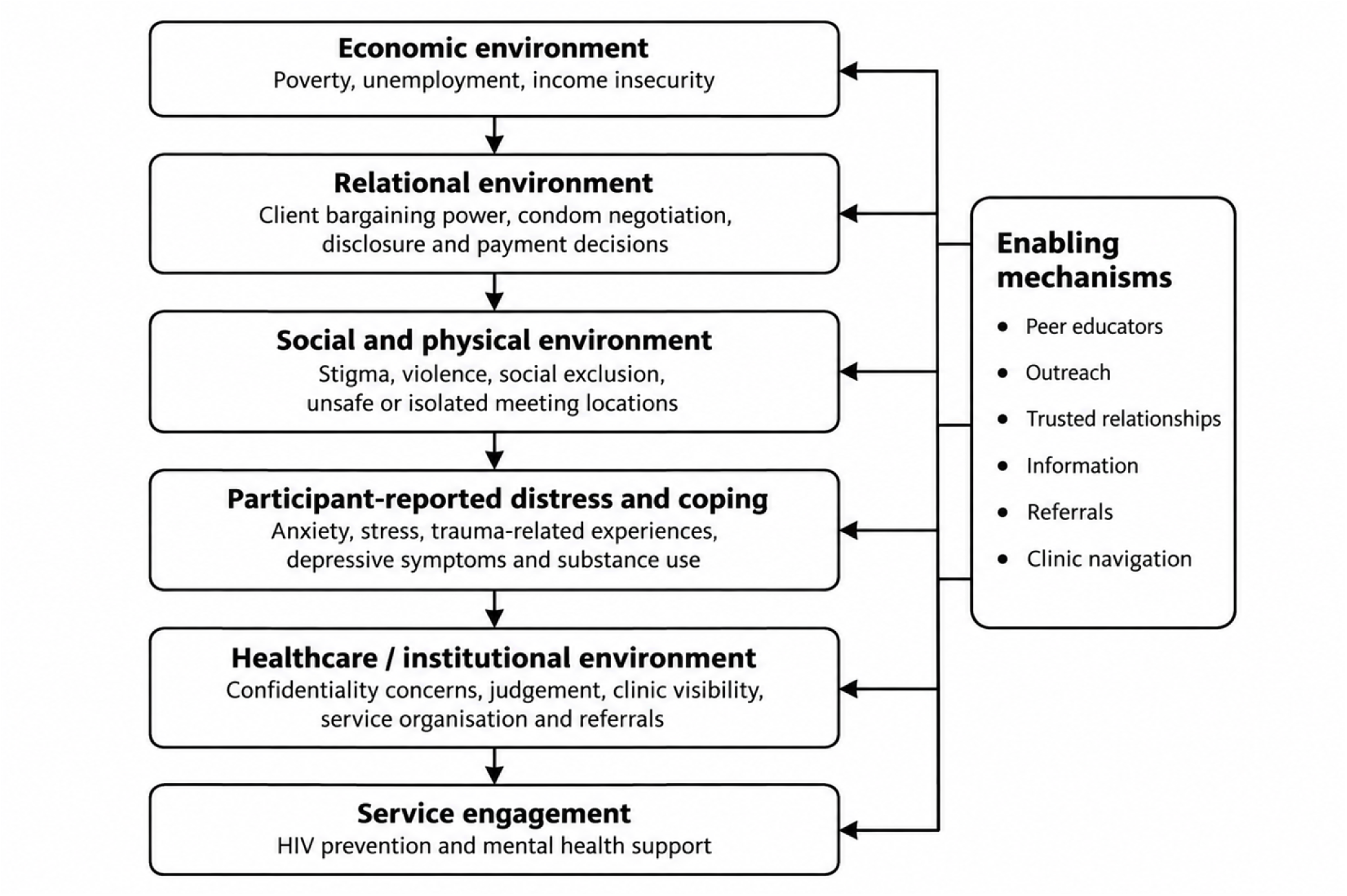
**Conceptual interpretation of participants’ accounts using the Risk Environment Framework.**

**Table 2.**
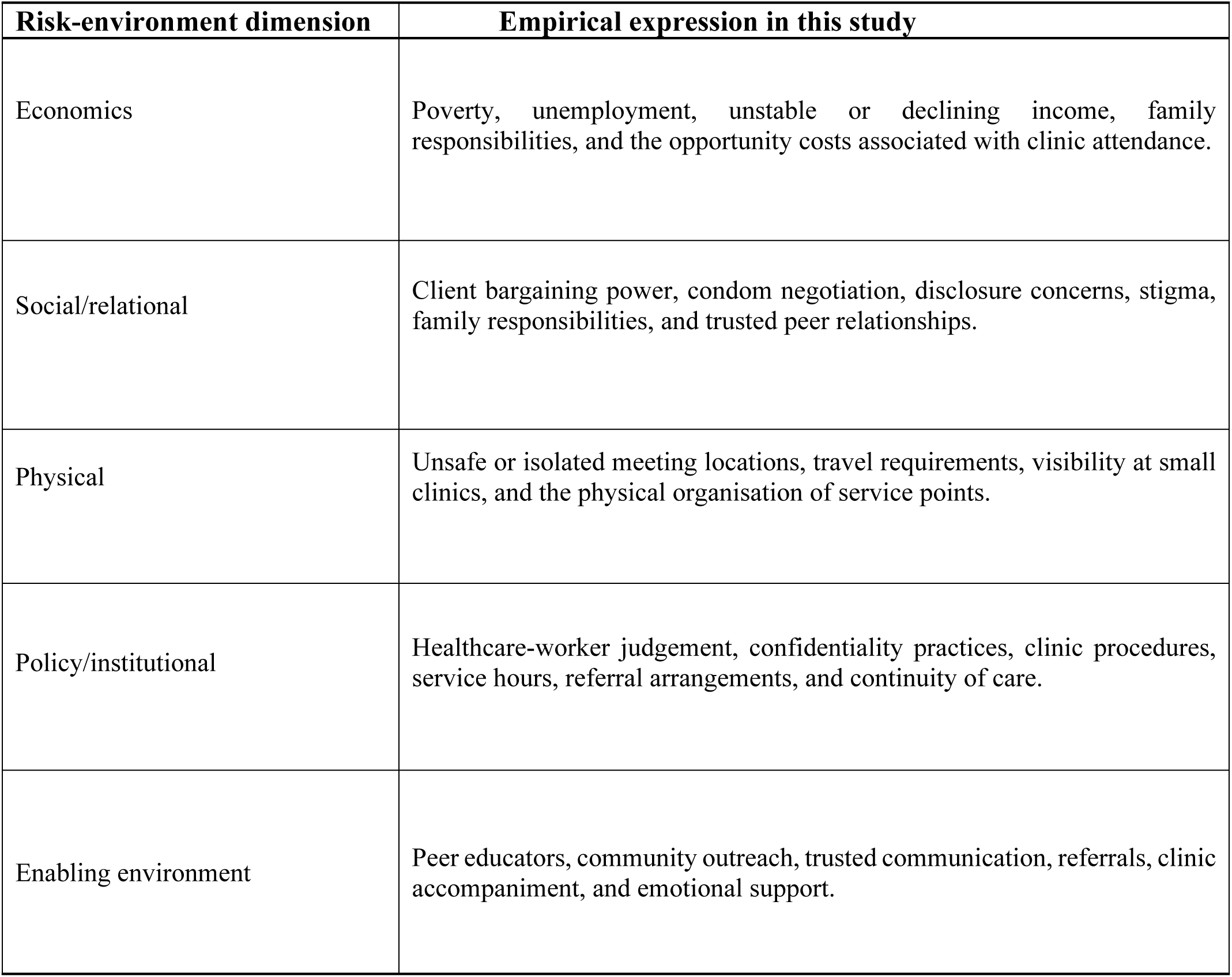
Analytical mapping of the findings onto the Risk Environment Framework.

| **Risk-environment dimension** | **Empirical expression in this study** |
| --- | --- |
| Economics | Poverty, unemployment, unstable or declining income, family responsibilities, and the opportunity costs associated with clinic attendance. |
| Social/relational | Client bargaining power, condom negotiation, disclosure concerns, stigma, family responsibilities, and trusted peer relationships. |
| Physical | Unsafe or isolated meeting locations, travel requirements, visibility at small clinics, and the physical organisation of service points. |
| Policy/institutional | Healthcare-worker judgement, confidentiality practices, clinic procedures, service hours, referral arrangements, and continuity of care. |
| Enabling environment | Peer educators, community outreach, trusted communication, referrals, clinic accompaniment, and emotional support. |

### Prevention Knowledge and Prevention Agency

The first analytical contribution is the distinction between prevention knowledge and prevention agency. Participants described knowledge of condoms, HIV testing and PrEP, yet they also described client payment, safety concerns and immediate income needs as part of decisions about whether preferred prevention practices could be implemented. This finding complements epidemiological evidence on the continuing HIV burden among female sex workers in South Africa (Kassanjee et al., 2022) and qualitative evidence that PrEP use can be motivated by self-protection, parenting and future aspirations (Makhakhe, Meyer-Weitz and Sliep, 2022). It also adds context to programme evidence showing that implementation strategies can improve PrEP persistence but do not eliminate wider structural constraints (Rao et al., 2023). Through the Risk Environment Framework, the findings suggest that prevention knowledge becomes actionable within, rather than outside, economic and relational conditions. The distinction is therefore between knowing what prevention options are available and having sufficient practical agency to use them consistently under conditions of unequal bargaining power.

Participants’ accounts of disclosure and condom negotiation further illustrate this distinction. Disclosure was weighed against possible rejection, stigma, violence or income loss, while condom use could become part of a negotiation over payment. These descriptions do not establish that economic insecurity directly causes unsafe sex, but they indicate how the perceived consequences of refusal may narrow participants’ practical choices. Rhodes (2002) emphasises that risk is produced through environments that distribute resources and power unevenly. The present study applies this principle to everyday sex-work encounters by showing how client relationships can mediate whether prevention preferences are feasible at a particular moment. The analytical contribution is therefore not that participants lacked agency, but that agency was exercised under conditions in which economic survival and interpersonal safety could compete with preferred health practices.

### Healthcare Access as Socially Produced

The second contribution is that healthcare access was experienced as socially and institutionally produced rather than as a simple question of physical availability. Participants described clinic visibility, anticipated judgement, confidentiality concerns and healthcare-worker interactions as factors that shaped whether services felt safe to use. This interpretation is consistent with Chen et al. (2022), who reported that HIV-related and sex work-related stigma is associated with poorer quality of life among female sex workers living with HIV. It also aligns with evidence from Comins et al. (2024) that structural vulnerabilities remain relevant to treatment coverage and viral suppression. International guidance calls for acceptable, non-discriminatory services for key populations (World Health Organization, 2022), while UNAIDS (2024) continues to identify stigma and discrimination as barriers to the HIV response. The West Rand findings add a district-level account of how these barriers can operate before, during and after a clinical encounter through fear of being seen, judged or involuntarily disclosed.

Peer accompaniment in participants’ accounts provides further evidence that access is relational. Some participants described formal services as easier to navigate when a trusted peer explained procedures, facilitated communication or accompanied them through a referral. This does not imply that peer accompaniment is necessary for every participant or that it caused successful care.

Rather, it suggests that trust and institutional navigation can alter how an existing service is experienced. The finding also exposes a limitation in defining access through service coverage alone: a service may exist and be geographically close while remaining difficult to use when its organisation produces fear of exposure or disrespect. Meaningful access therefore requires attention to how clinic systems, confidentiality practices and provider interactions shape the social experience of seeking care.

### Mental Health within the HIV Risk Environment

The third contribution is the positioning of participant-reported mental health distress within the HIV risk environment rather than as a separate programme concern. Participants described anxiety, stress, trauma-related experiences, depressive symptoms, mistrust and substance use alongside client negotiation, non-payment, unsafe locations and healthcare stigma. These reports should not be interpreted as clinical diagnoses, but they resonate with South African evidence documenting depression and syndemic factors among female sex workers living with HIV (Bhardwaj et al., 2023) and substance-use vulnerabilities among female sex workers in Soweto and Klerksdorp (Yeo et al., 2022). The present findings add detail on how participants themselves connected emotional distress with specific work and service experiences. They therefore support attention to mental health within HIV and primary healthcare pathways, while not establishing a causal effect of distress on service utilisation or demonstrating the effectiveness of any mental health intervention.

This interpretation supports the policy relevance of integrated care while remaining cautious about intervention claims. South Africa’s National Mental Health Policy Framework and Strategic Plan promotes integration of mental healthcare into community and district-level services (National Department of Health, 2023). Conteh, Latona and Mahomed (2023) found growing evidence for integrating mental health within HIV programmes, and Azuma et al. (2025) similarly report potential effectiveness of integrated HIV and mental health service delivery across low-and middle-income settings. The present study did not test an integrated model, and it therefore cannot conclude that a particular intervention would improve outcomes in the West Rand. Its contribution is to show that participants themselves did not experience emotional distress, HIV prevention, client power and healthcare engagement as neatly separable domains, providing a qualitative rationale for evaluating integrated approaches in future district-level research.

### Peer Educators as Boundary-Spanning Actors

The fourth contribution concerns peer educators as boundary-spanning actors between sex-worker communities and formal health services. Participants described peers as sources of understandable prevention information, emotional reassurance, referrals and clinic navigation. Community-led and peer-supported approaches are consistent with international guidance on key-population HIV services (UNAIDS, 2024; World Health Organization, 2022) and with South Africa’s emphasis on differentiated, community-centred responses (South African National AIDS Council, 2023). A recent scoping review similarly identifies peer education as a mechanism for supporting sexual-health knowledge and service linkage among female sex workers (Navidi et al., 2026). The West Rand findings extend this literature by emphasising the relational work peers performed: participants valued perceived absence of judgement, familiarity with sex-work realities and support in communicating with formal providers. Because recruitment also involved peer and service networks, however, these positive accounts should be interpreted alongside the possibility that participants with less favourable experiences of peer programmes were less likely to enter the study.

At the same time, the accounts indicate that peer support should complement rather than substitute for accessible and respectful formal services. Reliance on peers can reduce immediate uncertainty, but it does not by itself change clinic confidentiality, provider attitudes or availability of professional mental health care. The study also included participants across different gender identities, sexual orientations and work settings, reinforcing the need for peer and formal systems to avoid assuming one standard profile of a sex worker. However, subgroup numbers were too small for robust comparison, and the findings do not establish that a particular model is more effective for any identity group. The key implication is that peer relationships may provide an enabling layer within the risk environment while structural responsibility for safe, confidential and competent care remains with the health system.

### Implications for Policy and Practice

The findings support several implications for district-level policy and practice. First, participants’ accounts of waiting time, repeated appointments and income loss suggest that flexible service delivery may reduce the opportunity costs associated with seeking care. Discreet access pathways, confidential collection points and respectful consultation processes may also respond to the visibility and disclosure concerns described in the study. These implications are consistent with the South African National Strategic Plan’s emphasis on differentiated services, stigma reduction and community participation (South African National AIDS Council, 2023) and with World Health Organization (2022) guidance on acceptable services for key populations. The findings also support attention to mental health within HIV and primary healthcare pathways because participants described emotional distress and coping as intertwined with work and healthcare experiences. These are implications supported by the accounts and policy context; the study did not measure the effectiveness of any specific service redesign.

Several programme considerations follow from, but extend beyond, the qualitative evidence and would require prospective evaluation. District services could assess the feasibility of integrating brief mental health screening, support and referral into HIV prevention and primary healthcare contacts, strengthening confidentiality practices, training staff in non-judgemental communication, and formalising peer referral and navigation roles. Outreach strategies could also be reviewed to determine whether online or privately based sex workers, men and transgender sex workers are being reached alongside more visible street-based populations. Peer educators may be well placed to support information, emotional reassurance and referral, but complex mental health needs require access to appropriately trained practitioners and functional referral systems. These proposals should be treated as programme options informed by the findings and existing policy guidance, not as interventions whose effectiveness was demonstrated by this study.

## STRENGTHS AND LIMITATIONS

A major strength of the study was its focus on sex workers’ own accounts of HIV prevention, healthcare access and mental health, treating participants as knowledgeable interpreters of healthcare services, client relationships, peer support and emotional distress. The combined use of individual interviews and focus groups generated complementary forms of evidence: interviews created private space for sensitive accounts, while focus groups captured shared meanings, collective experiences and disagreement. The sample also included different gender identities, sexual orientations and work arrangements, which helped challenge a single assumed profile of sex work. The analysis was theoretically informed through Rhodes’ Risk Environment Framework and methodologically aligned with reflexive thematic analysis, including explicit recognition of researcher interpretation rather than consensus coding. Detailed reporting of context, recruitment, ethics, reflexivity and analytic procedures further supports assessment of the study’s credibility and transferability.

Several limitations should be considered. The study was conducted in one district, so the findings cannot be statistically generalised to all sex workers in South Africa and may reflect local economic conditions, service organisation, policing, migration and community attitudes. Recruitment through peer educators, outreach services and community networks may have favoured participants already connected to services and may have underrepresented sex workers who were more isolated from peer programmes. This is particularly relevant to the favourable accounts of peer educators. The individual interviews were relatively short at approximately 20– 30 minutes, which may have limited the depth available on some sensitive topics. Focus-group participants may have moderated responses because of social desirability or concerns that information could be repeated outside the group, and complete confidentiality between participants could not be guaranteed. Recall bias may also have affected accounts of previous healthcare, client and emotional experiences.

Further limitations arise from interpretation and representation. Translation into English may have resulted in some loss of nuance even though translated transcripts were cross-checked against audio recordings. The first author’s professional identity and interaction with participants may have influenced questioning and interpretation despite reflexive procedures. Formal participant member checking was not conducted, so findings were not returned to participants for confirmation. Small numbers of men, bisexual participants and transgender women limited subgroup analysis and increased the need to avoid deductive disclosure; consequently, the study cannot make robust comparative claims across gender identity or sexual orientation. The study also provides stronger evidence about participant-reported mental health distress than about the uptake, quality or outcomes of formal mental health support services; this distinction limits interpretation of the term “mental health support” and is one reason the manuscript frames the findings around mental health rather than service effectiveness. Finally, the qualitative design did not estimate prevalence, establish causal relationships or evaluate the effectiveness of peer-led, integrated mental health or differentiated service interventions.

## CONCLUSION

The study found that sex workers in the West Rand described HIV prevention, healthcare access and mental health within an interconnected economic, relational, physical and institutional environment. Participants generally demonstrated knowledge of prevention, but their accounts indicated that condom use, PrEP, disclosure and clinic attendance were negotiated alongside income needs, client demands, safety concerns, stigma and confidentiality. Participants also reported anxiety, stress, trauma-related experiences, depressive symptoms, mistrust and substance use in relation to everyday work and healthcare encounters. These accounts are not evidence of clinically diagnosed disorders or direct causal relationships. Rather, they show why mental health and HIV prevention cannot be understood solely at the level of individual knowledge or intention.

Peer educators were described as trusted bridges to care through information, emotional reassurance, referrals and clinic navigation. Their role was best understood as an enabling feature of the wider risk environment rather than as a substitute for formal healthcare. The findings support consideration of discreet access pathways, strengthened confidentiality, integrated mental health support and referral, and service approaches capable of reaching sex workers across different work settings and identities. These are areas for programme development and prospective evaluation, not interventions whose effectiveness was established by this study. The central contribution is the demonstration of how prevention knowledge, practical agency, healthcare access, participant-reported distress and peer support intersect in everyday experience.

## Data Availability

The qualitative data are not publicly available because the recordings and transcripts contain sensitive information concerning sex work, HIV prevention, mental health, stigma, service use and personal experiences. De-identified excerpts may be made available by the corresponding author following a reasonable request, subject to research ethics approval, participant confidentiality requirements and applicable data-protection procedures.

## DATA AVAILABILITY STATEMENT

The underlying qualitative recordings and full transcripts are not publicly available because they contain sensitive and potentially identifiable information concerning sex work, HIV status and prevention, mental health experiences, stigma, healthcare use, violence and client interactions. Public release would be inconsistent with the confidentiality commitments made to participants and the conditions under which ethical approval was granted. Requests from qualified researchers for access to a restricted, de-identified analytic dataset or selected de-identified excerpts may be considered in consultation with the Sefako Makgatho Health Sciences University Research Ethics Committee (SMUREC) and the corresponding author. Requests should describe the proposed research purpose, the specific data required, evidence of relevant ethics approval where applicable, and arrangements for secure data management. Any access would be subject to SMUREC requirements, participant confidentiality protections, applicable South African data-protection requirements and any permissions required by the West Rand Health District or relevant organisational gatekeepers. Unrestricted access to raw audio files or identifiable transcripts will not be provided.

## FUNDING

This study was supported for doctorial study under postgraduate scholarship/ Partial Cost of Study (PCS) from National Research Foundation (NRF) PMDS250604319356 of South Africa. The funder had no role in study design, data collection, analysis and decisions to prepare and publish the manuscript.

## ACKNOWLEDGEMENTS

The authors acknowledge the sex workers who participated in the study and shared their experiences. Appreciation is extended to the peer educators, community-based organisations, healthcare workers and district stakeholders who supported recruitment, referrals and fieldwork.

## Notes

### Competing Interest Statement

The authors have declared no competing interest.

### Author Declarations

Ethical approval was obtained from the Sefako Makgatho Health Sciences University Research Ethics Committee (SMUREC), South Africa (Reference No. SMUREC/H/544/2026:PG). Permission to conduct the study was also obtained from the West Rand Health District (Reference No. GP_202510_091).

## REFERENCES

Azuma, H., Yeh, P.T., Kennedy, C.E., Fonner, V.A., O’Reilly, K.R. and Sweat, M.D. (2025) ’Integration of mental health and HIV service delivery in low- and middle-income countries: A systematic review of effectiveness’, Cambridge Prisms: Global Mental Health, 12, e112. doi: 10.1017/gmh.2025.10066.

Bhardwaj, A., Comins, C.A., Guddera, V., Mcingana, M., Young, K., Phetlhu, R., et al. (2023) ‘Prevalence of depression, syndemic factors and their impact on viral suppression among female sex workers living with HIV in eThekwini, South Africa’, BMC Women’s Health, 23, 232. doi: 10.1186/s12905-023-02392-2.

Braun, V. and Clarke, V. (2021) ‘One size fits all? What counts as quality practice in (reflexive) thematic analysis?’, Qualitative Research in Psychology, 18(3), pp. 328–352. doi: 10.1080/14780887.2020.1769238.

Chen, C., Baral, S., Comins, C.A., Mcingana, M., Wang, L., Phetlhu, D.R., et al. (2022) ‘HIV-and sex work-related stigmas and quality of life of female sex workers living with HIV in South Africa: A cross-sectional study’, BMC Infectious Diseases, 22, 910. doi: 10.1186/s12879-022-07892-4.

Comins, C.A., Baral, S., Mcingana, M., Shipp, L., Phetlhu, D.R., Young, K., Guddera, V., Hausler, H. and Schwartz, S. (2024) ‘ART coverage and viral suppression among female sex workers living with HIV in eThekwini, South Africa: Baseline findings from the Siyaphambili study’, PLOS Global Public Health, 4(5), e0002783. doi: 10.1371/journal.pgph.0002783.

Conteh, N.K., Latona, A. and Mahomed, O. (2023) ‘Mapping the effectiveness of integrating mental health in HIV programs: A scoping review’, BMC Health Services Research, 23, 396. doi: 10.1186/s12913-023-09359-x.

Creswell, J.W. and Creswell, J.D. (2023) Research Design: Qualitative, Quantitative, and Mixed Methods Approaches. 6th edn. Thousand Oaks, CA: SAGE.

Hennink, M. and Kaiser, B.N. (2022) ‘Sample sizes for saturation in qualitative research: A systematic review of empirical tests’, Social Science & Medicine, 292, 114523. doi: 10.1016/j.socscimed.2021.114523.

Kassanjee, R., Welte, A., Otwombe, K., Jaffer, M., Milovanovic, M., Hlongwane, K., et al. (2022) ‘HIV incidence estimation among female sex workers in South Africa: A multiple methods analysis of cross-sectional survey data’, The Lancet HIV, 9(11), pp. e781–e790. doi: 10.1016/S2352-3018(22)00201-6.

Kiger, M.E. and Varpio, L. (2020) ‘Thematic analysis of qualitative data: AMEE Guide No. 131’, Medical Teacher, 42(8), pp. 846–854. doi: 10.1080/0142159X.2020.1755030.

Makhakhe, N.F., Meyer-Weitz, A. and Sliep, Y. (2022) ‘Motivating factors associated with oral pre-exposure prophylaxis use among female sex workers in South Africa’, Journal of Health Psychology, 27(12), pp. 2820–2833. doi: 10.1177/13591053211072674.

National Department of Health (2023) National Mental Health Policy Framework and Strategic Plan 2023-2030. Pretoria: National Department of Health.

Navidi, I., Shakibazadeh, E., Akbarpour, S. and Raisi, F. (2026) ‘Peer-education strategies for enhancing sexual health services among female sex workers: A scoping review’, BMC Women’s Health, 26, 40. doi: 10.1186/s12905-025-04208-x.

O’Brien, B.C., Harris, I.B., Beckman, T.J., Reed, D.A. and Cook, D.A. (2014) ’Standards for reporting qualitative research: A synthesis of recommendations’, Academic Medicine, 89(9), pp. 1245–1251. doi: 10.1097/ACM.0000000000000388.

Rao, A., Mhlophe, H., Pretorius, A., Mcingana, M., McLoughlin, J., Shipp, L., Baral, S., Hausler, H., Schwartz, S. and Lesko, C. (2023) ‘Effect of implementation strategies on pre-exposure prophylaxis persistence among female sex workers in South Africa: An interrupted time series study’, The Lancet HIV, 10(12), pp. e807–e815. doi: 10.1016/S2352-3018(23)00262-X.

Rhodes, T. (2002) ‘The risk environment: A framework for understanding and reducing drug-related harm’, International Journal of Drug Policy, 13(2), pp. 85–94. doi: 10.1016/S0955-3959(02)00007-5.

Shannon, K., Crago, A.L., Baral, S.D., Bekker, L.G., Kerrigan, D., Decker, M.R., Poteat, T., Wirtz, A.L., Weir, B., Boily, M.C., Butler, J., Strathdee, S.A. and Beyrer, C. (2018) ‘The global response and unmet actions for HIV and sex workers’, The Lancet, 392(10148), pp. 698–710. doi: 10.1016/S0140-6736(18)31439-9.

South African National AIDS Council (2023) National Strategic Plan for HIV, TB and STIs 2023-2028. Pretoria: SANAC.

Tong, A., Sainsbury, P. and Craig, J. (2007) ‘Consolidated criteria for reporting qualitative research (COREQ): A 32-item checklist for interviews and focus groups’, International Journal for Quality in Health Care, 19(6), pp. 349–357. doi: 10.1093/intqhc/mzm042.

UNAIDS (2024) HIV and Sex Workers: Thematic Briefing Note. Geneva: Joint United Nations Programme on HIV/AIDS.

World Health Organization (2022) Consolidated Guidelines on HIV, Viral Hepatitis and STI Prevention, Diagnosis, Treatment and Care for Key Populations. Geneva: World Health Organization.

Yeo, E.J., Hlongwane, K., Otwombe, K., Hopkins, K.L., Variava, E., Martinson, N., Strathdee, S.A., Coetzee, J. and Milovanovic, M. (2022) ‘Key risk factors for substance use among female sex workers in Soweto and Klerksdorp, South Africa: A cross-sectional study’, PLOS ONE, 17(1), e0261855. doi: 10.1371/journal.pone.0261855.

